# Different treatment durations of loperamide in preventing pyrotinib-induced diarrhea: A randomized, parallel-group sub-study of the phase II PHAEDRA trial

**DOI:** 10.1101/2024.08.19.24311958

**Authors:** Changjun Wang, Yan Lin, Ying Xu, Feng Mao, Jinghong Guan, Xuejing Wang, Yanna Zhang, Xiaohui Zhang, Songjie Shen, Ying Zhong, Bo Pan, Li Peng, Xin Huang, Xi Cao, Ru Yao, Xintong Zhou, Zecheng He, Yuhan Liu, Jie Lang, Chenggang Li, Yidong Zhou, Qiang Sun

## Abstract

**Background:** Pyrotinib, a pan-HER tyrosine kinase inhibitor, demonstrates efficacy in the treatment of HER2-positive breast cancer. However, the frequent occurrence of treatment-emergent diarrhea necessitating discontinuation, impacts patient outcomes.

**Methods:** In this multicenter, open-label, phase II PHAEDRA study enrolling early stage HER2-positive patients for postoperative treatment with nab-paclitaxel and pyrotinib, 120 patients were included in a sub-study and randomly divided into two groups to receive 21 days and 42 days of loperamide for primary prophylaxis of diarrhea, followed by as-needed usage. The primary outcome was the incidence of grade ≥3 diarrhea.

**Results:** Fifty-eight patients in the 21-day group and 59 patients in the 42-day group received at least one dose of pyrotinib. With a median follow-up of 12.1 months, all patients experienced diarrhea of any grade, with grade ≥3 events in 39.7% of the 21-day group and 42.4% of the 42-day group (relative risk: 0.94; 95% confidence interval: 0.61-1.45). The most common treatment-emergent adverse events, other than diarrhea, were hypoesthesia, vomiting, nausea, and rash, mostly grade 1-2, except for one patient with a grade ≥3 decreased neutrophil count in each group.

**Conclusion:** No significant differences were observed between 21-day and 42-day loperamide durations in preventing grade ≥3 diarrhea. Considering the economic cost and patient compliance, 21-day loperamide prophylaxis might represent a more pragmatic and appropriate approach for clinical application.

**Trial registration:** ClinicalTrials.gov, NCT04659499

## Introduction

Pyrotinib is a pan-human epidermal growth factor receptor (HER) tyrosine kinase inhibitor (TKI) targeting HER1, HER2, and HER4 independently developed in China. Since its introduction, pyrotinib has demonstrated significant survival benefits in patients with HER2-positive breast cancer. In the phase III PHENIX trial, pyrotinib in combination with capecitabine achieved a median progression-free survival (PFS) of 11.1 months for HER2-positive metastatic breast cancer, showing a risk reduction of 82% for disease progression or death compared with the control group (placebo plus capecitabine).^1^ Similarly, in the phase III PHOEBE trial, pyrotinib combined with capecitabine obtained a median PFS of 12.5 months in HER2-positive metastatic breast cancer, reducing the risk of disease progression or death by 61% compared with lapatinib combined with capecitabine.^2^ In the PHILA trial, pyrotinib in combination with trastuzumab and docetaxel had a median PFS of 24.3 months in the first-line treatment of HER2-positive metastatic breast cancer, demonstrating significant superiority compared to trastuzumab combined with docetaxel plus placebo (median PFS=10.4 months).^3^ The phase III PHEDRA study on the neoadjuvant treatment of early stage HER2-positive breast cancer, revealed a significant improvement in the total pathologic complete response (tpCR) rate with the combination of pyrotinib, trastuzumab, and docetaxel beyond that of placebo, trastuzumab, and docetaxel (41% vs. 22%, p <0.0001).^4^

However, similar to other TKIs, diarrhea remains a significant adverse event frequently occurring during pyrotinib treatment, with an incidence rate exceeding 95% in these studies.^1–4^ Specifically, the incidences of grade ≥3 treatment-emergent diarrhea were 30.8% and 30.5% in the PHENIX and PHOEBE studies with the treatment of pyrotinib plus capecitabine,^1, 2^ respectively, and an even more pronounced 46.5% in the PHILA study and 44.4% in the PHEDRA study with the treatment of pyrotinib combined with trastuzumab plus docetaxel.^3, 4^ Severe diarrhea can necessitate discontinuation of pyrotinib, subsequently impacting its anti-tumor efficacy, posing a clinical safety concern that warrants attention.

In the CONTROL study, the cohort receiving loperamide as a primary prophylaxis for diarrhea over the initial 56 days documented a grade ≥3 diarrhea incidence rate of 31% during neratinib administration.^5^ In contrast, the ExteNET study, which did not employ any diarrhea prophylactic strategies, reported a 40% rate of grade ≥3 diarrhea.^6^ Consequently, the Chinese Society of Clinical Oncology (CSCO) guideline recommends the prophylactic use of loperamide starting with the first dose of neratinib, continuing for 56 days.^7^ In the NALA study, a 21-day loperamide of primary prophylaxis similarly reduced the incidence of grade ≥3 diarrhea during combined neratinib and capecitabine therapy to 24.4%.^8^

Currently, there are no clinical studies on the prevention of pyrotinib-induced diarrhea. For patients whose pyrotinib therapy was interrupted due to diarrhea, the CSCO guidelines suggest a 21-day prophylactic treatment with loperamide upon re-initiating pyrotinib. To explore the feasibility of loperamide’s primary prophylaxis for pyrotinib-induced diarrhea and to further optimize diarrhea prophylactic strategies, this study evaluated the incidence of diarrhea in patients with early stage HER2-positive breast cancer receiving albumin-bound paclitaxel combined with pyrotinib, employing both 42-day and 21-day primary prophylaxis with loperamide.

## Methods

### Study design and patients

This was a sub-study of the multicenter, open-label, single-arm phase II PHAEDRA study (NCT04659499) designed to investigate different prophylactic strategies for treatment-emergent diarrhea that occurred during pyrotinib treatment. The study was approved by the institutional ethics committee of Peking Union Medical College Hospital (approval number: HS-2617) and was conducted in accordance with the 2008 Declaration of Helsinki. The PHAEDRA study included patients with tumors ≤3 cm, lymph node-negative (N0) or micro-metastatic (N1mi), HER2 positive breast cancer from Peking Union Medical College Hospital, Beijing Longfu Hospital, Weihai Municipal Hospital, and Weihai Maternal and Child Health Hospital in China to receive adjuvant treatment with nab-paclitaxel and pyrotinib within 90 days after tumor resection. In this sub-study, 120 patients from the main study who signed the sub-study informed consent were 1:1 randomly assigned to two groups to receive 21-days (the 21-day group) and 42-days (the 42-day group) of loperamide as primary prophylaxis for treatment-emergent diarrhea (Figure 1). The full eligibility criteria for the main study and this sub-study have been previously reported.^9^

**Figure 1.**
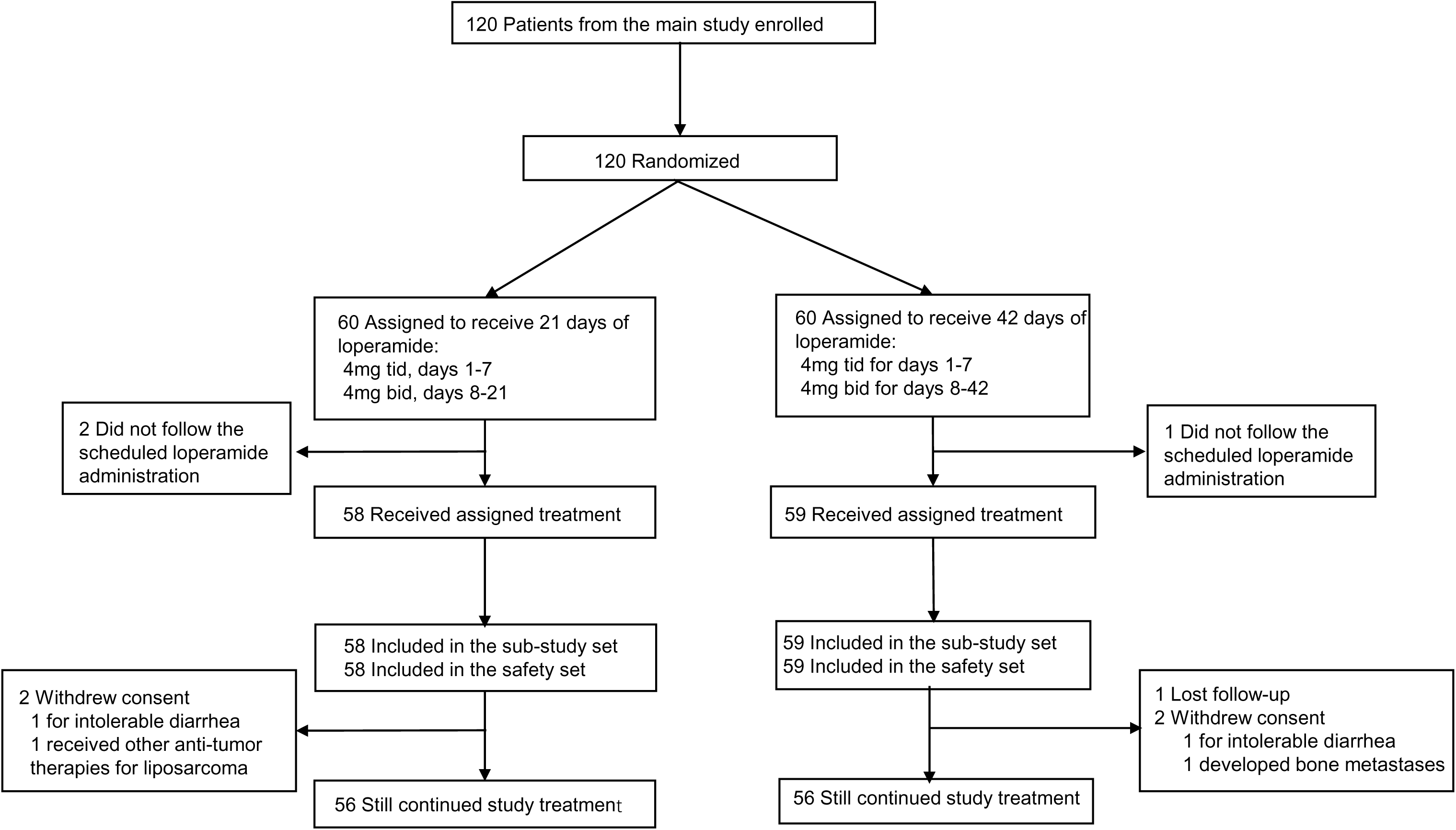
Patient flow.

### Treatment procedure

All patients were administered intravenous nab-paclitaxel (260 mg/m^2^ once every 21-days cycle for 4 cycles and oral pyrotinib (400 mg) once daily for 1 year as adjuvant therapy. For primary prophylaxis of treatment-emergent diarrhea, the 21-day group additionally received loperamide during cycle 1 of adjuvant therapy at a dose of 4 mg three times a day on days 1–7 and 4 mg twice a day on days 8–21; the 42-day group received loperamide during cycles 1 and 2 of adjuvant therapy at a dose of 4 mg three times a day on days 1–7 and 4 mg twice a day on days 8–42. Once the primary prophylaxis was completed, loperamide was administered as needed for treatment-emergent diarrhea management during the remaining treatment duration with pyrotinib. When grade 1–2 treatment-emergent diarrhea with complications (including but not limited to abdominal cramping, grade ≥2 nausea/vomiting, fever, pyemia, decreased neutrophil count, bleeding, and dehydration) or grade 3 treatment-emergent diarrhea occurs, pyrotinib treatment is interrupted and resumed until recovery to grade 0–1 without complications. Dose reduction is not required after recovery for the first time, but should be reduced to 320 mg at the next time and to 240 mg at most. If the continuous interruption time was more than 28 days, patients were withdrawn from the study. When grade 4 treatment-emergent diarrhea occurs, pyrotinib is permanently discontinued. Patients were assessed at the study center every 3 weeks ±7 days for the initial 10 weeks and subsequently every 3 months ±14 days until a full year of pyrotinib treatment was completed. Follow-up continued for 28 days after the last dose of pyrotinib was given.

### Outcomes

The primary endpoint of the sub-study was the incidence of grade ≥3 treatment-emergent diarrhea from the first dose to 28 days after the last dose of the study drug.

The secondary endpoints of this study included the incidence and severity of treatment-emergent diarrhea during the first two cycles (42 days) of adjuvant therapy; the incidence and severity of constipation; the onset time, frequency, and duration of grade ≥3 treatment-emergent diarrhea; the relationship between treatment-emergent diarrhea and study drugs; the incidence of dose reduction, discontinuation, and hospitalization due to treatment-emergent diarrhea; all treatment-emergent adverse events (TEAEs) during the study duration; and the quality of life (QoL) questionnaire score using the Functional Assessment of Cancer Therapy-Breast (FACT-B). The severity of treatment-emergent diarrhea was graded for consistency with the National Cancer Institute Common Terminology Criteria for Adverse Events (NCI CTCAE), version 5.0, assuming that the subjects had one stool per day at baseline.

### Statistical analysis

All the safety analyses were descriptive. Kaplan–Meier curves are shown for the percentage of subjects in each group who did not develop diarrhea of any grade, and for those who did not have diarrhea of grade ≥3. The two groups were compared using the log-rank test. QoL analyses were carried out in all patients, including the safety population who underwent baseline and one post-baseline QoL assessment. A mixed-model for repeated measures (MMRM) was used to analyze the data. The mean (standard error) of the observed scores over time was calculated. Statistical analyses were performed using R (version 4.1.2).

## Results

### Baseline characteristics

Between Jane 8, 2021, and November 22, 2022, 120 patients completed enrollment and randomization. Three patients did not follow the scheduled loperamide administration, and 117 patients received at least one dose of pyrotinib: 58 in the 21-day group and 59 in the 42-day group. All of them were included in the sub-study analysis set and the safety analysis set. At the cut-off date on September 27, 2023, two patients withdrew from the 21-day group for intolerable diarrhea and received other anti-tumor therapies for liposarcoma on the face. In the 42-day group, 1 patient was lost to follow-up and 2 withdrew from the study for intolerable diarrhea and bone metastases. The baseline characteristics are summarized in Table 1. The median follow-up time was 12.1 months (interquartile range [IQR]: 9.5, 12.8).

**Table 1.** Baseline characteristics.

| **Characteristics** | **21-day**<br>**loperamide**<br>**(N=58)** | **42-day**<br>**loperamide**<br>**(N=59)** |
| --- | --- | --- |
| **Age (years), median (IQR)** | 50 (27, 72) | 52 (29, 68) |
| **Female, n (%)** | 58 (100) | 59 (100) |
| **ECOG performance status, n (%)** |  |  |
| 0 | 29 (50.0) | 37 (62.7) |
| 1 | 29 (50.0) | 22 (37.3) |
| **Hormone receptor status, n (%)** |  |  |
| ER+ and/or PR+ | 40 (69.0) | 33 (55.9) |
| ER- and PR- | 18 (31.0) | 26 (44.1) |
| **Stage, n (%)** |  |  |
| IA | 49 (84.5) | 47 (79.7) |
| IB | 3 (5.2) | 1 (1.7) |
| IIA | 6 (10.3) | 11 (18.6) |
| **Menopausal, n (%)** | 25 (43.1) | 30 (50.8) |

IQR, interquartile range; ECOG, Eastern Cooperative Oncology Group; ER, estrogen receptor; PR, progesterone receptor.

### Treatment-emergent diarrhea

All 117 patients developed treatment-emergent diarrhea (Table 2). At 1 year and 28 days after the first dose of pyrotinib, 76 episodes of grade ≥3 treatment-emergent diarrhea occurred in 23 patients (39.7%) in the 21-day group and 96 episodes occurred in 24 patients (40.7%) in the 42-day group. The relative risk for developing grade ≥3 treatment-emergent diarrhea was 0.97 (95% confidence interval [CI]: 0.63, 1.52). The median number of episodes for each patient who developed grade ≥3 treatment-emergent diarrhea was 2 (IQR: 1, 5) and 1 (IQR: 1, 3) in groups A and B, respectively. The median duration per grade ≥3 treatment-emergent diarrhea episodes was 2 (IQR: 2, 2) days for the 21-day group and 2 (IQR: 2, 3) days for the 42-day group. In the 21-day group, the median cumulative duration of grade ≥3 treatment-emergent diarrhea per patient, accounting for the total duration of all individual episodes, was 5 days (IQR: 2, 12). Meanwhile, the 42-day group had a median of 2.5 days (IQR: 2, 6.8). Most cases of grade ≥3 diarrhea developed in the first 3 months (Figure 2). In the 21-day group, the median time to the first occurrence of grade ≥3 diarrhea was 9 days (IQR: 7, 22) after the initial dose of pyrotinib. In contrast, for the 42-day group, this event was observed at a median time of 16 days (IQR: 7.3, 75.3). In the first 42 days, 43 episodes of grade ≥3 treatment-emergent diarrhea occurred in 21 patients (36.2%) in the 21-day group with each episode lasting 2 days (IQR: 2, 3), whereas 32 episodes occurred in 16 patients (27.1%) in the 42-day group, with each episode lasting 2 days (IQR: 2, 3).

**Figure 2.**
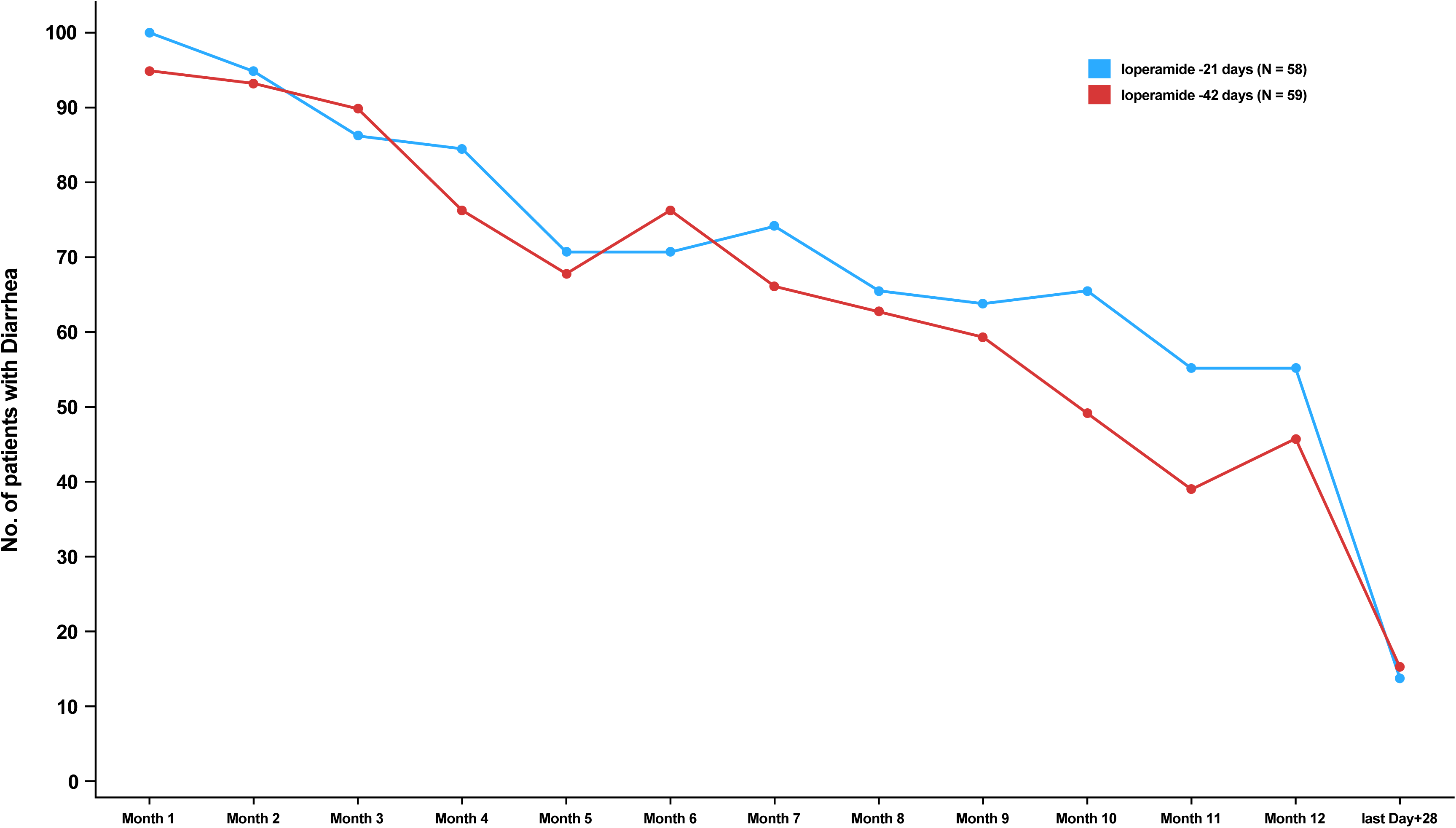
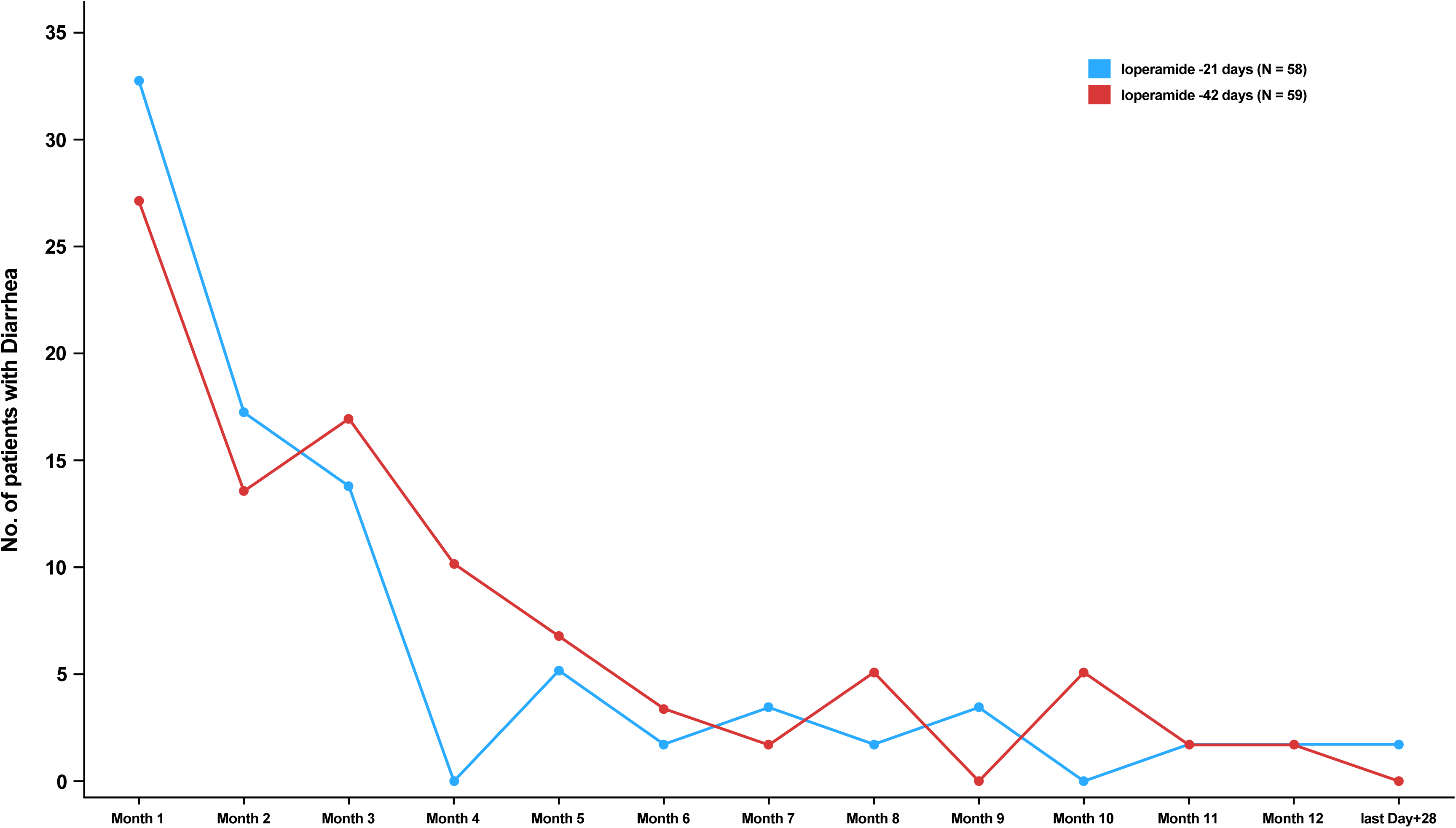
Incidence of diarrhea of any grade (a) or grade ≥3 (b) by month.

**Table 2.**
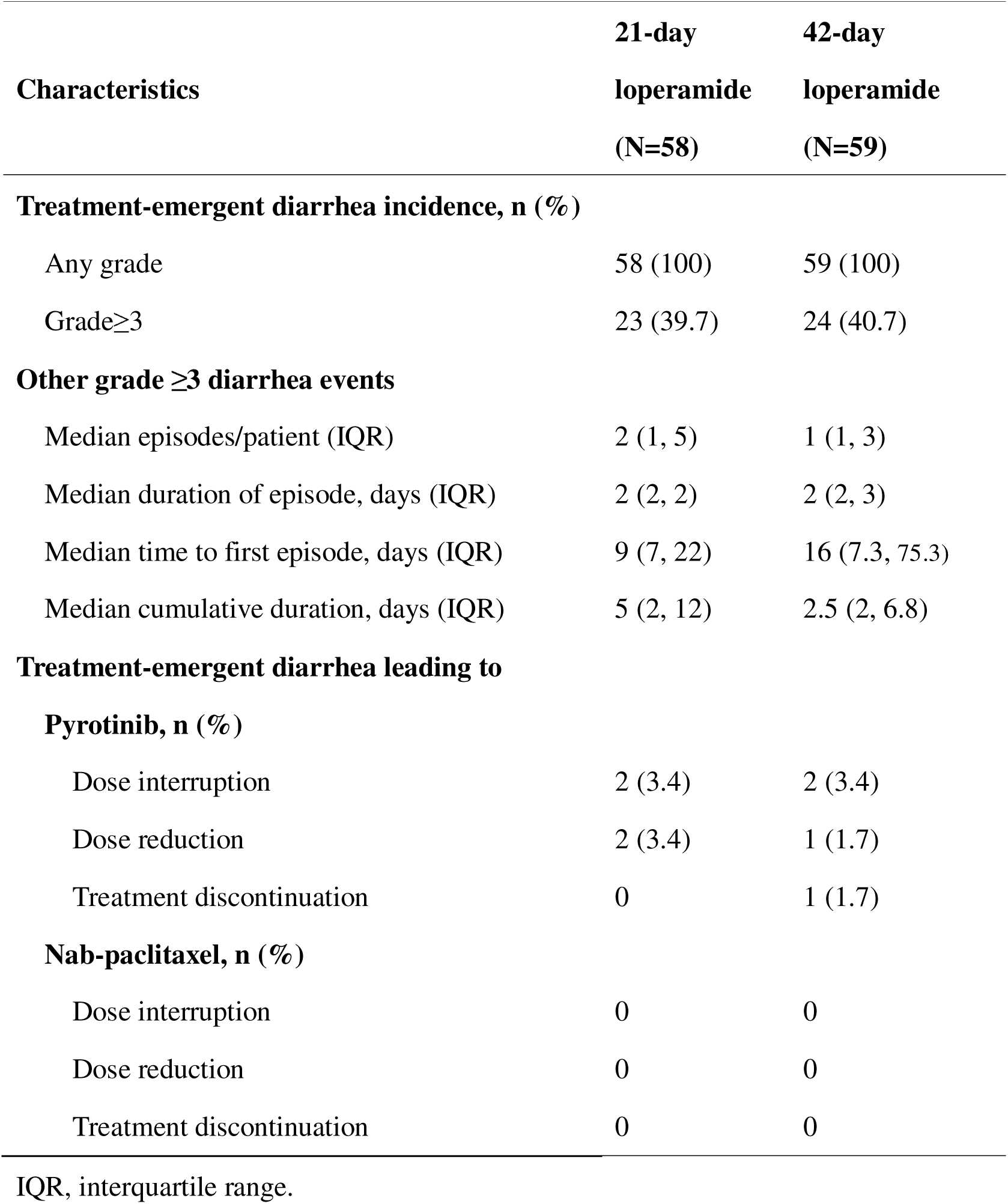
Characteristics of treatment-emergent diarrhea.

From the Kaplan–Meier curves, 42-days loperamide prophylaxis did not reduce the proportion of patients who developed treatment-emergent diarrhea of any grade (Figure 3a) or grade ≥3 (Figure 3b).

**Figure 3.**
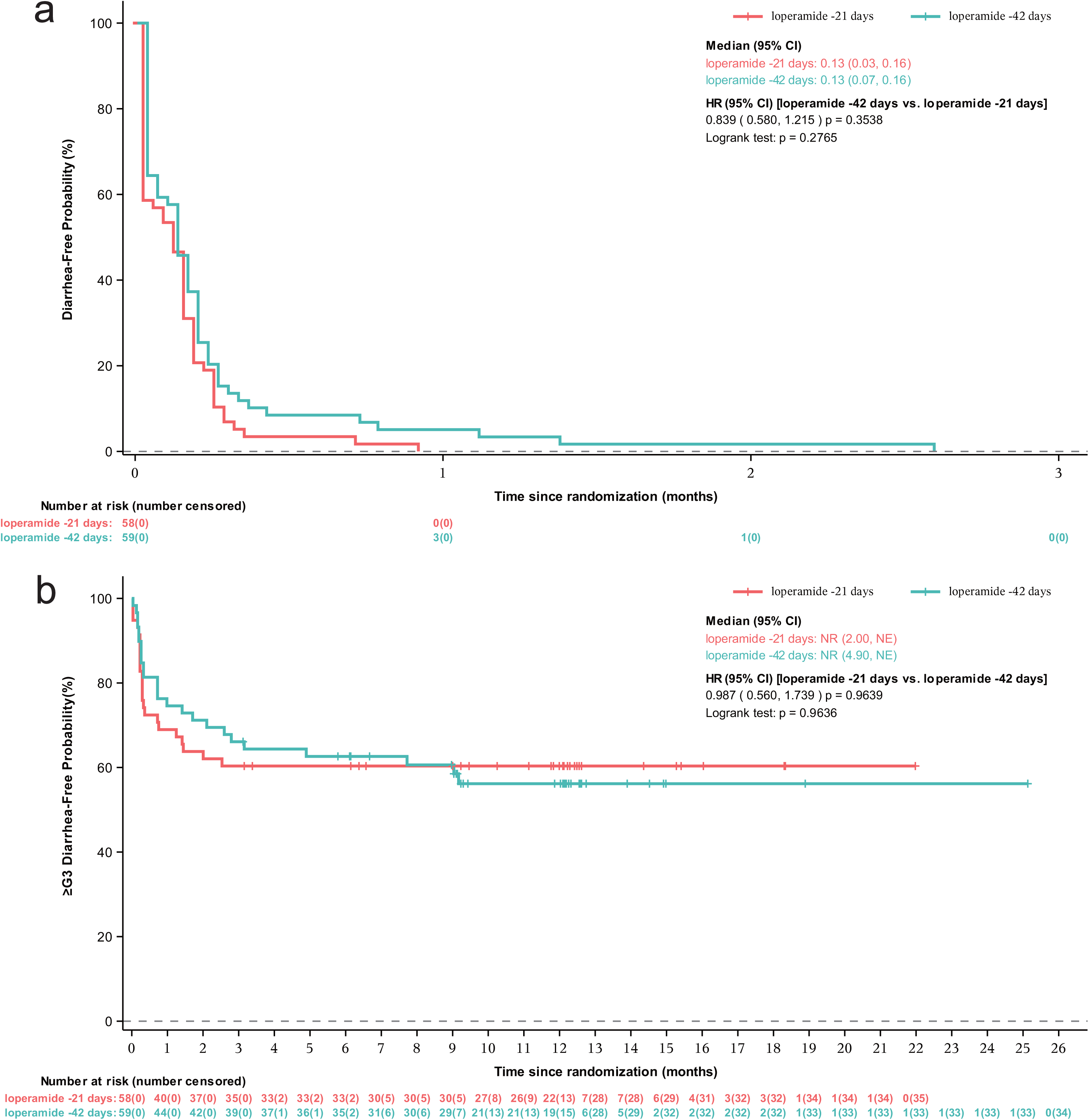
Percentage of patients who did not develop a diarrhea of any grade (a) or grade ≥3 (b) after pyrotinib administration.

One patient in both groups discontinued pyrotinib treatment for intolerant diarrhea. The proportion of patients interrupting pyrotinib treatment due to diarrhea was 3.4% in each group (Table 2). Pyrotinib was discontinued in one patient with diarrhea in the 42-day group, while there were no discontinuations in the 21-day group. Two patients in the 21-day group and one patient in the 42-day group required a dose reduction of pyrotinib due to diarrhea. Despite the high incidence of diarrhea in the first two cycles of treatment, only one patient (1.7%) in the 21-day group interrupted pyrotinib, and there was no pyrotinib discontinuation or nab-paclitaxel reduction or discontinuation, probably due to continuous administration of loperamide.

In the 21-day group, the incidences of treatment-related diarrhea of any grade and grade ≥3 during the combination treatment of nab-paclitaxel plus pyrotinib in the first 12 weeks were 91.4% and 34.5%, respectively, whereas those of pyrotinib alone were 94.8% and 13.8%, respectively. In the 42-day group, treatment-related diarrhea of any grade occurred in 88.1% of patients during combination treatment versus 94.9% during pyrotinib monotherapy. Furthermore, 28.8% of patients during combination treatment and 25.4% of patients during pyrotinib monotherapy experienced treatment-related diarrhea of grade ≥3.

### Non-diarrhea TEAEs

The most common TEAEs, other than diarrhea, were hypoesthesia, vomiting, nausea, and rash in both groups. The safety profile in this study was similar to that in previous reports on pyrotinib and nab-paclitaxel (Table 3). Most TEAEs were of grade 1-2. However, in each group, there was one case (1.7%) of grade ≥3 neutrophil count decrease, and one case of grade ≥3 white blood cell count decrease. Only 1 patient across the groups required granulocyte colony-stimulating factor administration to manage these adverse events. In one patient, the administration of nab-paclitaxel and pyrotinib was modified with dose reductions due to hepatic function abnormalities. Four patients (6.9%) in the 21-day group and five (8.5%) in the 42-day group developed grade 1 constipation during the primary prophylaxis period, leading to a reduction of loperamide in three patients. Intestinal obstruction or more serious sequelae of constipation have not yet been reported. No serious TEAEs have been reported. No deaths occurred.

**Table 3.**
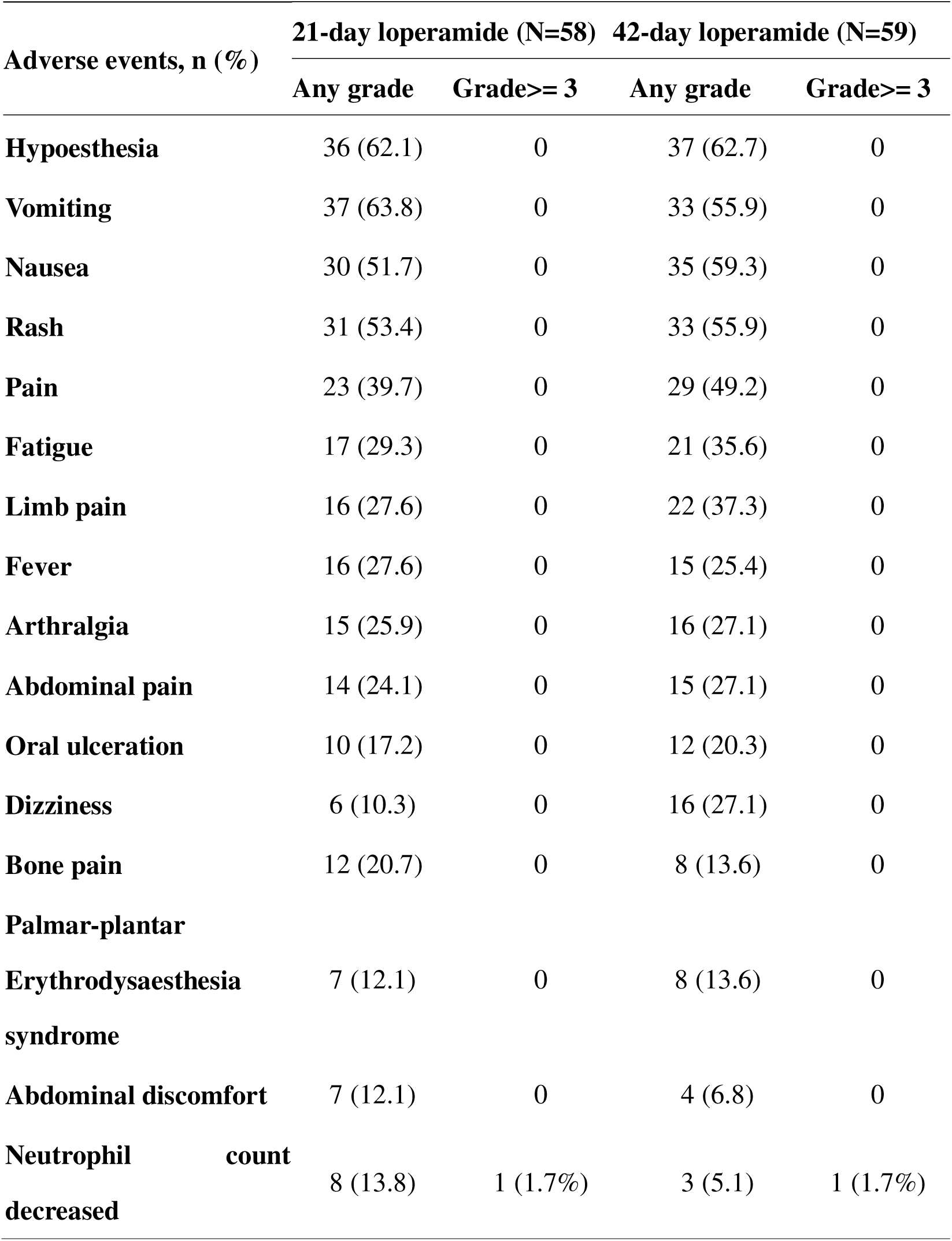
Treatment-emergent non-diarrhea adverse events with an incidence of >10%.

### QoL

Based on the designated follow-up regimen, QoL assessment utilizing the FACT-B questionnaire was administered tri-weekly for the initial 3-month period and subsequently on a quarterly basis thereafter. FACT-B scores were analyzed using MMRM model, and the results showed that in both the 21-day group and the 42-day group, a discernible decline from baseline in FACT-B aggregate scores was observed during the preliminary two months (Figure 4). This decline was consistent with a clinically significant difference (7-8 points). However, a resurgence in scores was evident from the third month. The score trajectories of the two groups displayed marked divergence in the ninth month. Notably, only 31 questionnaires were collected in the 12^th^ month.

**Figure 4.**
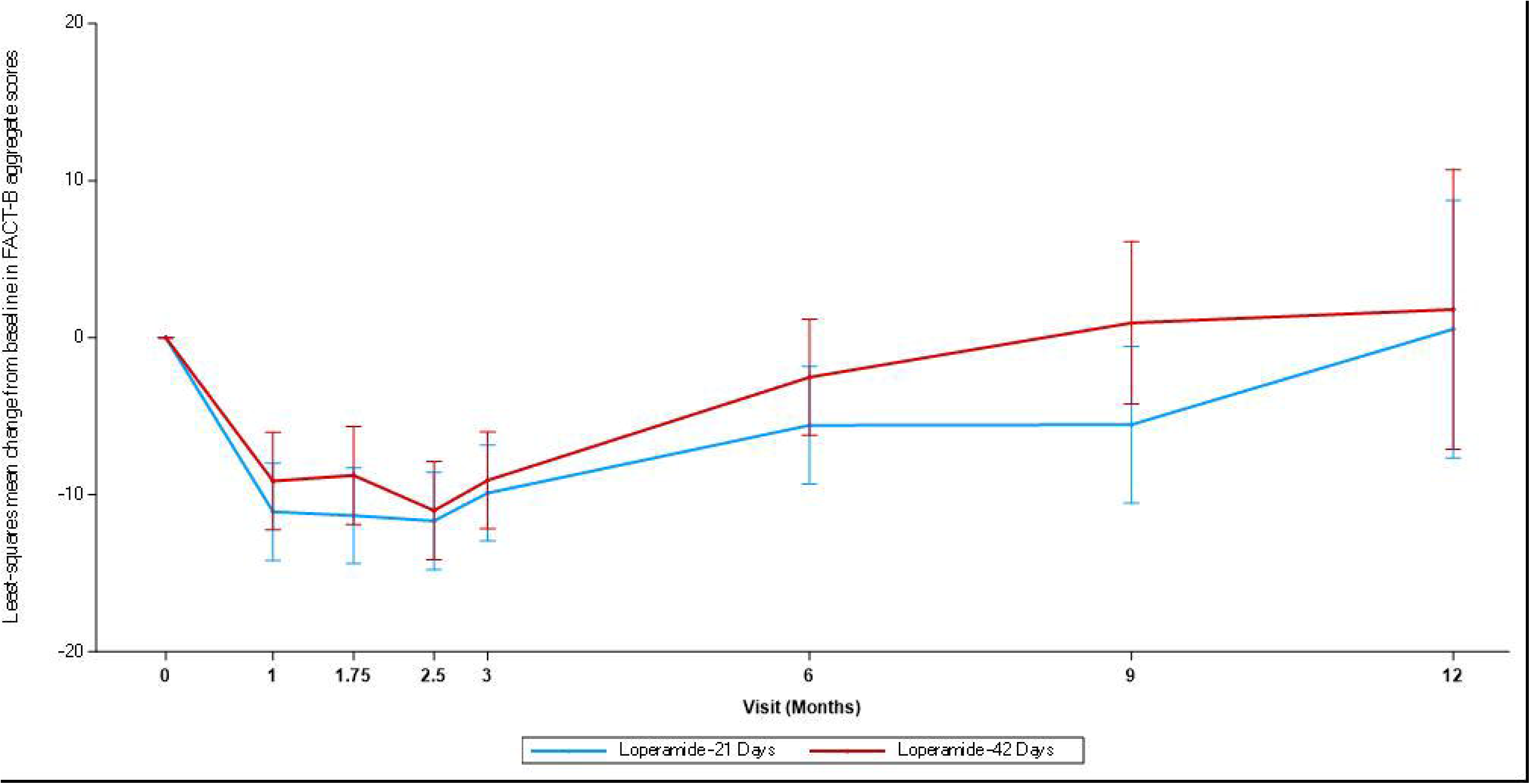
Mean change from baseline in Functional Assessment of Cancer Therapy (Breast) total scores.

## Discussion

The ErbB receptor plays a pivotal role in the maintenance of intestinal barrier integrity. Inhibition of the epidermal growth factor receptor (EGFR) results in diminished proliferation of intestinal mucosal epithelial cells, augmented apoptosis, and heightened endoplasmic reticulum stress. This compromises the healing capacity of the intestinal epithelium, leading to mucosal atrophy and impaired intestinal absorption. Furthermore, both HER2 and EGFR are expressed on the membranes of intestinal epithelial cells. Their synergistic action modulates chloride ion secretion through the phosphoinositide 3-kinase and protein kinase C pathways. Upon inhibition of these receptors, there is an excess of chloride ions in the intestinal lumen, which reduces water absorption and predisposes the patient to aqueous diarrhea. Additional mechanisms contributing to TKI-induced diarrhea include inflammatory responses,^10^ aberrant bile acid secretion,^11^ and dysbiosis of the intestinal microbiota. Given these mechanisms, diarrhea is inevitable and is one of the most frequent and significant TEAEs during TKIs treatment.

The incidence of diarrhea varies among TKIs. For lapatinib, the probability of diarrhea ranges from 45% to 79.7%, with grade 3 or higher diarrhea occurring in 6% to 20.7% of cases.^12–14^ In contrast, the incidence of diarrhea during neratinib treatment reaches up to 95.3%, with grade 3 or higher observed in 40% of cases.^6^ Pyrotinib has a diarrhea incidence rate similar to neratinib, ranging between 94.8% and 98.4%, with approximately 31% of these cases being grade 3 or above.^1, 2^ Thus, the CSCO guideline, referencing the CONTROL study which investigated prophylaxis strategies for neratinib-induced diarrhea, recommends that for patients who discontinued pyrotinib due to diarrhea, a 21-day prophylactic treatment with loperamide should be administered once pyrotinib re-initiates. As grade 3 diarrhea predominantly occurs during the first cycle of pyrotinib administration,^2, 15^ it is hypothesized that using loperamide as primary prophylaxis during the initial 21 days might mitigate the onset of pyrotinib-induced diarrhea. Nonetheless, a notable incidence of grade 3 diarrhea was still observed during the second cycle.^2^ In the PHEDRA study, prophylactic diarrhea management with loperamide reduced the incidence of grade 3 diarrhea to 36.1% (26/72) compared with 50.0% (53/106) in those without such management. Specifically, there was a 15.1% reduction during the first cycle (29.2% vs. 44.3%) and an 11.7% reduction in the second cycle (10.1% vs. 21.8%).^4^ In this context, our study explored two regimens of loperamide: a 21-day and a 42-day course, initiated concurrently with the first dose of pyrotinib, as primary prophylaxis for diarrhea. The results indicated no significant differences between the 21-day and 42-day prophylaxis regimens in terms of overall diarrhea incidence (100% vs.100%) or the occurrence of grade 3 or higher diarrhea (39.7% vs. 42.4%). Achieving a balance between constipation and diarrhea has been a clinical challenge when using loperamide to manage TKI-induced diarrhea. However, in this study, the incidence of constipation remained low for both the 21-day (6.9%) and 42-day (8.5%) loperamide regimens, with a severity of only grade 1.

Severe diarrheal symptoms can prompt dose reduction or discontinuation of pyrotinib, which may potentially affect therapeutic outcomes. In the PHOEBE study, one patient (1%) discontinued pyrotinib due to diarrhea.^2^ Meanwhile, in another phase II study, diarrhea did not result in pyrotinib discontinuation, but it did lead to dose reductions in two patients (3%).^15^ In the present study, both the 21-day and 42-day loperamide groups had only one patient (1.7%) who discontinued treatment. In each group, two patients (3.4%) interrupted pyrotinib, while dose reductions were observed in two patients (3.4%) in the 21-day group and one case (1.7%) in the 42-day group.

Frequent episodes of diarrhea can compromise a patient’s quality of life and affect medication adherence. In the current study, there was a decline in the patients’ FACT-B scores during the initial two months. This decrease is potentially attributable to the early onset of diarrhea, commonly observed with the concurrent administration of nab-paclitaxel and pyrotinib during the early stages of treatment. As the treatment proceeded, the quality-of-life scores gradually improved.

In this study, a combination of pyrotinib and nab-paclitaxel was used as an anti-tumor therapeutic regimen. These TEAEs were consistent with previously reported profiles associated with both pyrotinib and albumin-bound paclitaxel. Given that nab-paclitaxel can also induce diarrhea, the frequency of diarrhea in this study, when co-administered with pyrotinib and nab-paclitaxel, was elevated compared to that in previous studies combining pyrotinib with capecitabine. The lower incidence of neutropenia compared with that reported in previous study involving nab-paclitaxel (34%), probably due to the fact that the breast cancer patients in that study were all in advanced stages with poorer overall health conditions, resulted in fewer use of leukocyte-raising agents. Notably, the hematological toxicity observed with the combination of nab-paclitaxel and anti-HER2 TKI in our study was lower than that observed with paclitaxel combined with an anti-HER2 antibody in the APT trial.

This study has several limitations. First, as the main study has currently completed patient enrollment, but is still ongoing, the data are not yet mature; therefore, there is a lack of patients who have not received prophylactic treatment for diarrhea to serve as a control group. Second, although the study incorporated randomization for group allocation, blinding was not employed. Finally, in the later stages of follow-up, numerous patients did not submit their quality-of-life questionnaires, rendering this portion of the scoring potentially imprecise.

In conclusion, no significant differences were observed between 21-day and 42-day loperamide durations in preventing grade ≥3 diarrhea. Considering the economic cost and patient compliance, 21-day loperamide prophylaxis might represent a more pragmatic and appropriate approach for clinical application.

## Data Availability

The raw clinical data were protected and not available owing to data privacy laws. The de-identified datasets supporting the findings of this study are available for academic purposes on request from the corresponding author, Qiang Sun, for five years, with the approval of the Institutional Ethical Committees.

## Acknowledgments

We would like to acknowledge the patients and their families, the study investigators, and the clinical site staff. We thank Xiaoyue Wu (Department of Medical Affairs, Jiangsu Hengrui Pharmaceuticals Co., Ltd.) for her input in study design and data interpretation, Yitao Wang (Department of Medical Affairs, Jiangsu Hengrui Pharmaceuticals Co., Ltd.) for statistical support, and Yilan Wu (Department of Medical Affairs, Jiangsu Hengrui Pharmaceuticals Co., Ltd.) for medical writing assistance.

## Ethics approval and consent to participate

The study was approved by the institutional ethics committee at the Peking Union Medical College Hospital (approval number: HS-2617) with informed consent obtained from all patients in accordance with the Declaration of Helsinki. This trial was registered at ClinicalTrials.gov, number NCT04659499.

## Consent for publication

Not applicable.

## Author contributions

We had full access to all data in the trial and took responsibility for the integrity of the data and accuracy of the data analysis. Y.D.Z. and Q.S. contributed to the conceptualization and design of the trial. C.J.W., Y.L., Y.X., F.M., J.H.G, X.J.W., Y.N.Z, X.H.Z., S.J.S., Y.Z., B.P., L.P., X.H., X.C., R.Y., X.T.Z, Z.C.H., Y.H.L, C.G.L., J.L. collected and assembled the data. C.J.W. and Y.L. performed the statistical analyses and wrote the manuscript. All the authors participated in writing the paper and approved the final version of the manuscript.

